## Supplemental Material for "Clinical gait analysis using video-based pose estimation: multiple perspectives, clinical populations, and measuring change"

**Fig. S1**

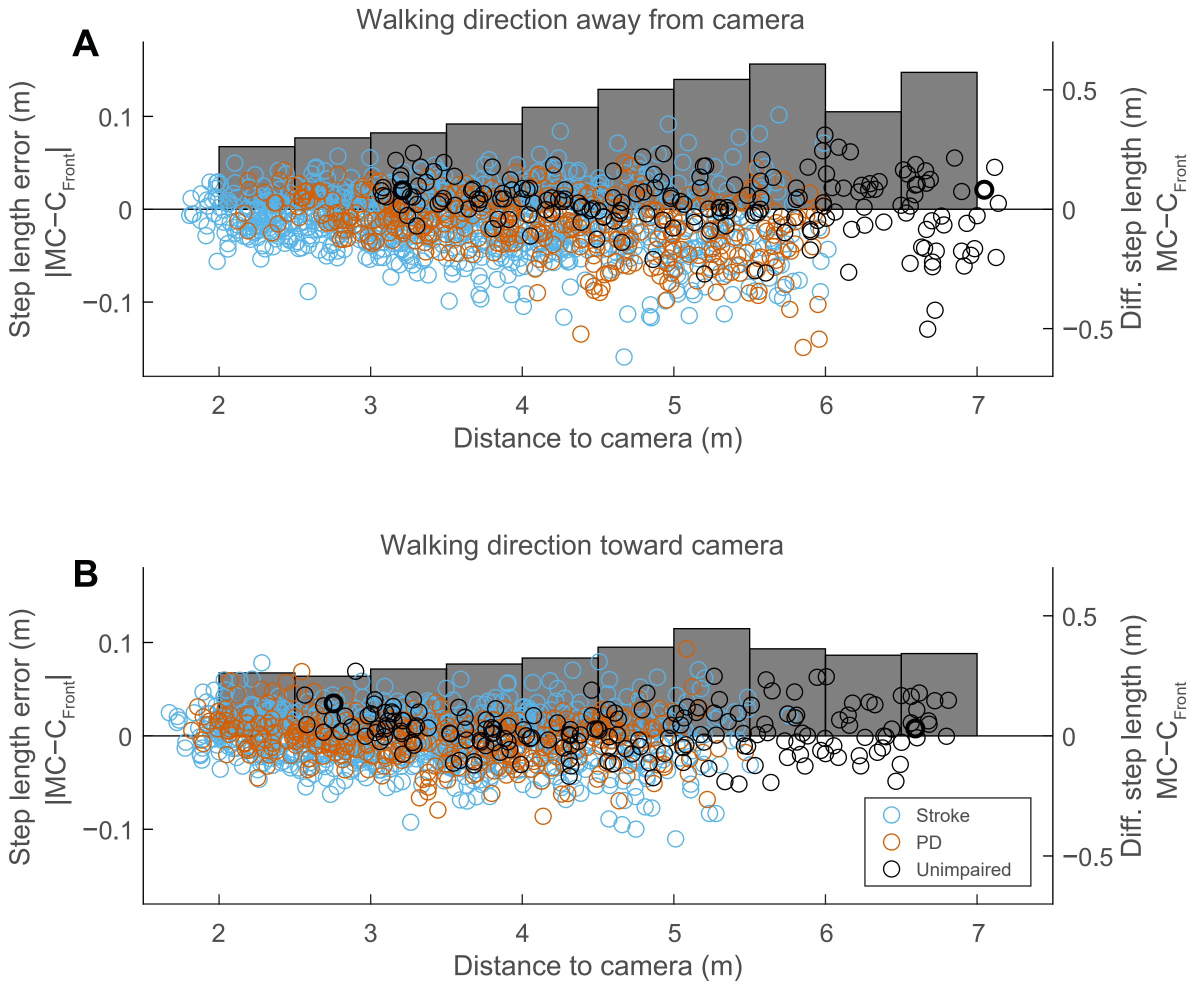


**Fig. S1. Step length errors and differences of frontal plane workflow relative to person's distance to camera.** Erorrs and differences relative to distance when the person is walking away from the camera (A) or toward the camera (B). Bar graphs show average step length errors binned across 0.5 m and values are represented on left-hand y-axes. Circles show step length differences for individual steps and values are shown on right-hand y-axes.

**Fig. S2**

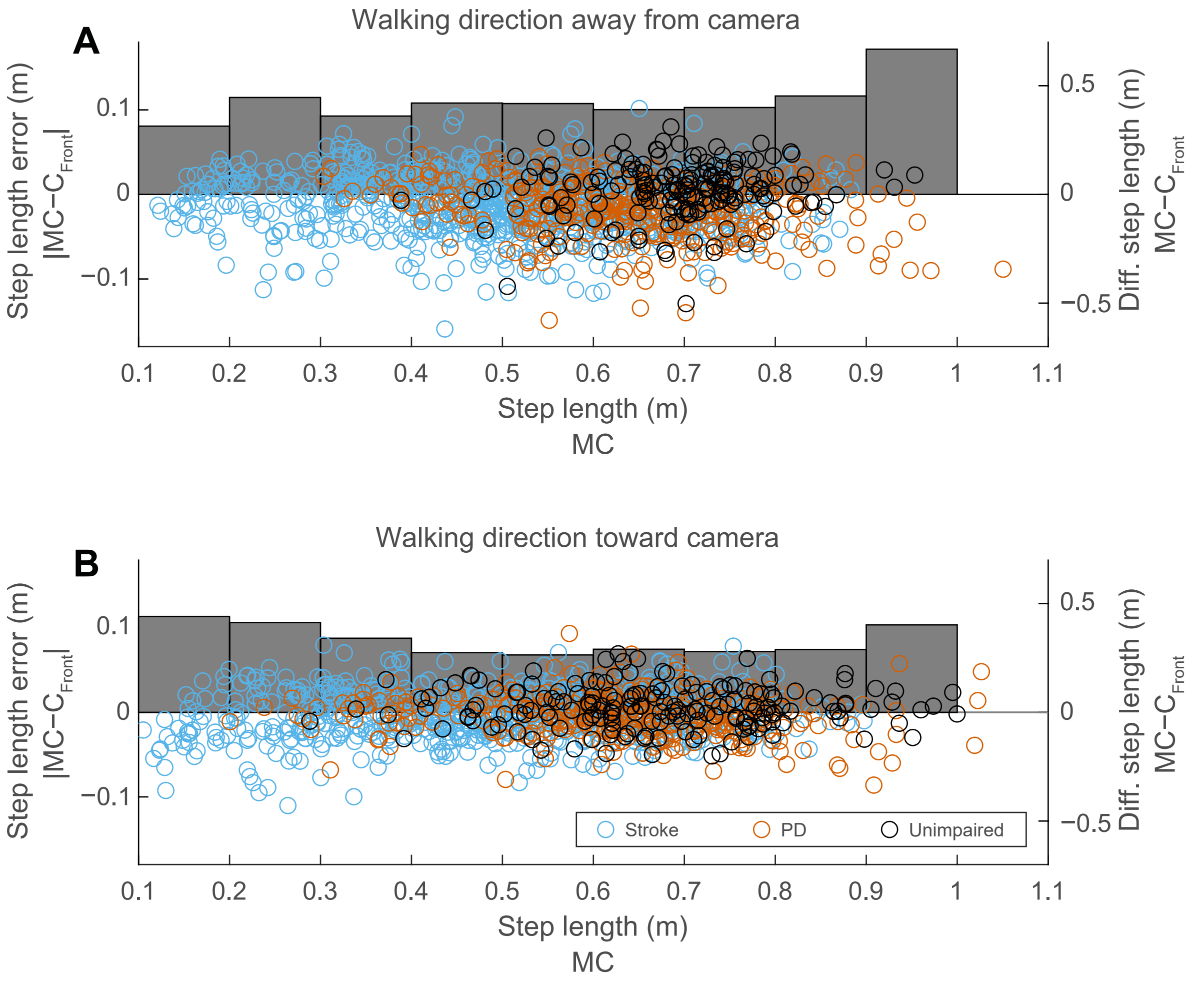


**Fig. S2. Step length errors and differences of frontal plane workflow relative to magnitude of step length.** Erorrs and differences relative to step length magnitude when the person is walking away from the camera (A) or toward the camera (B). Bar graphs show average errors binned across 0.1-m intervals and values are represented on left-hand y-axes. Data points show differences for individual steps and values correspond to right-hand y-axes.

#### Fig. S3

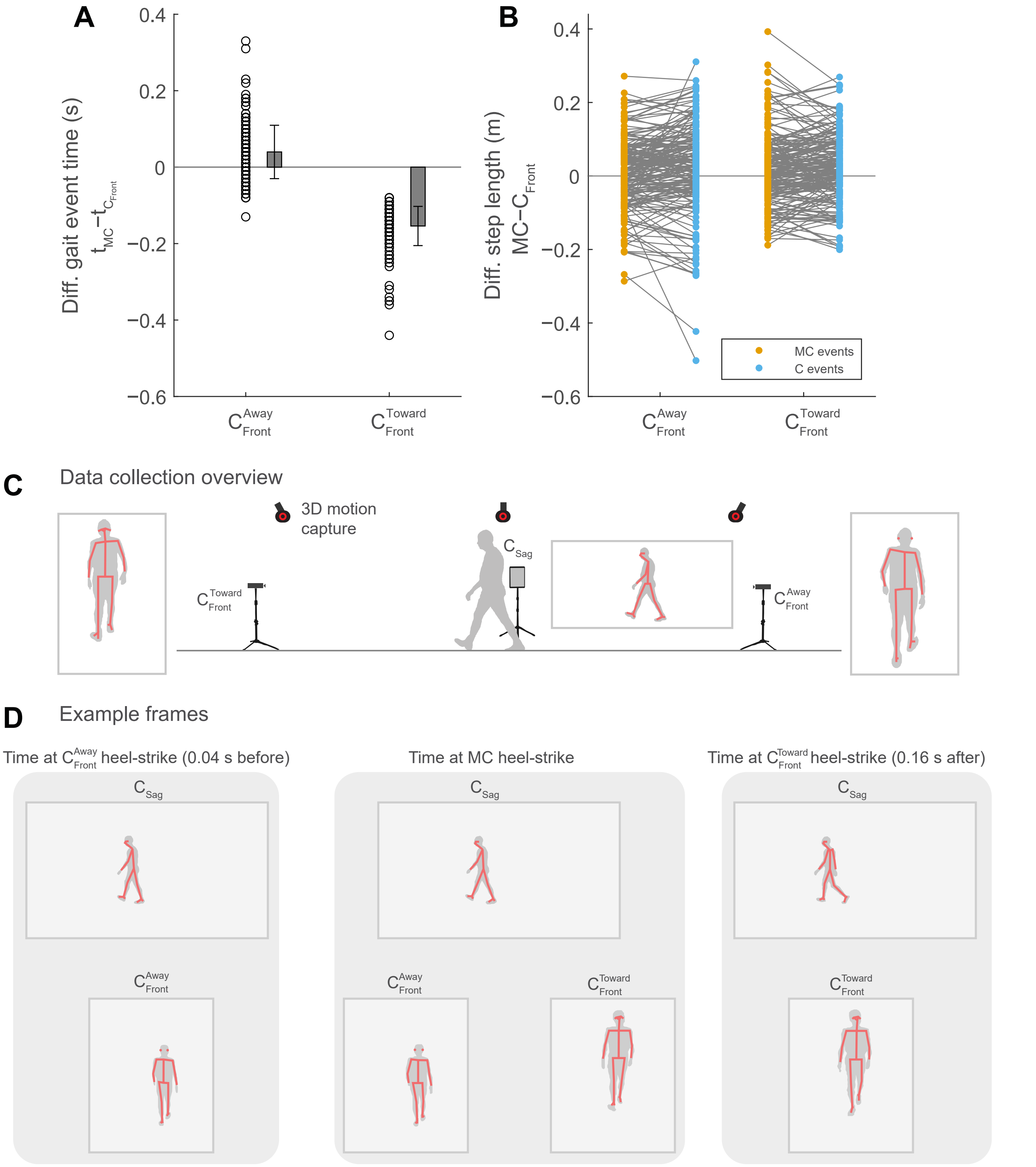


**Fig. S3. Influence of gait event timings on step length errors when using frontal plane workflow.** Gait events times (heel-strikes) found using the frontal plane workflow are on average detected 0.04 s before the gait events detected from motion capture when the person is walking away from the frontal plane camera, and 0.15 s after motion capture when the person is walking toward the frontal plane camera (A). The effect of differences in gait event times on step length differences is shown in panel B. Data collection overview (C). Example frames of sagittal and frontal camera views of one participant at gait event times detected by the frontal camera that the person walks away from (left), detected by motion capture (middle) and detected by the frontal camera that the person walks toward (right). Note that this analysis can only be performed using the unimpaired data set because recordings of motion capture and video data where synchronized. Note that photographs in panels C and D have been replaced by silhouettes to conform to medRxiv policy.

### Fig. S4

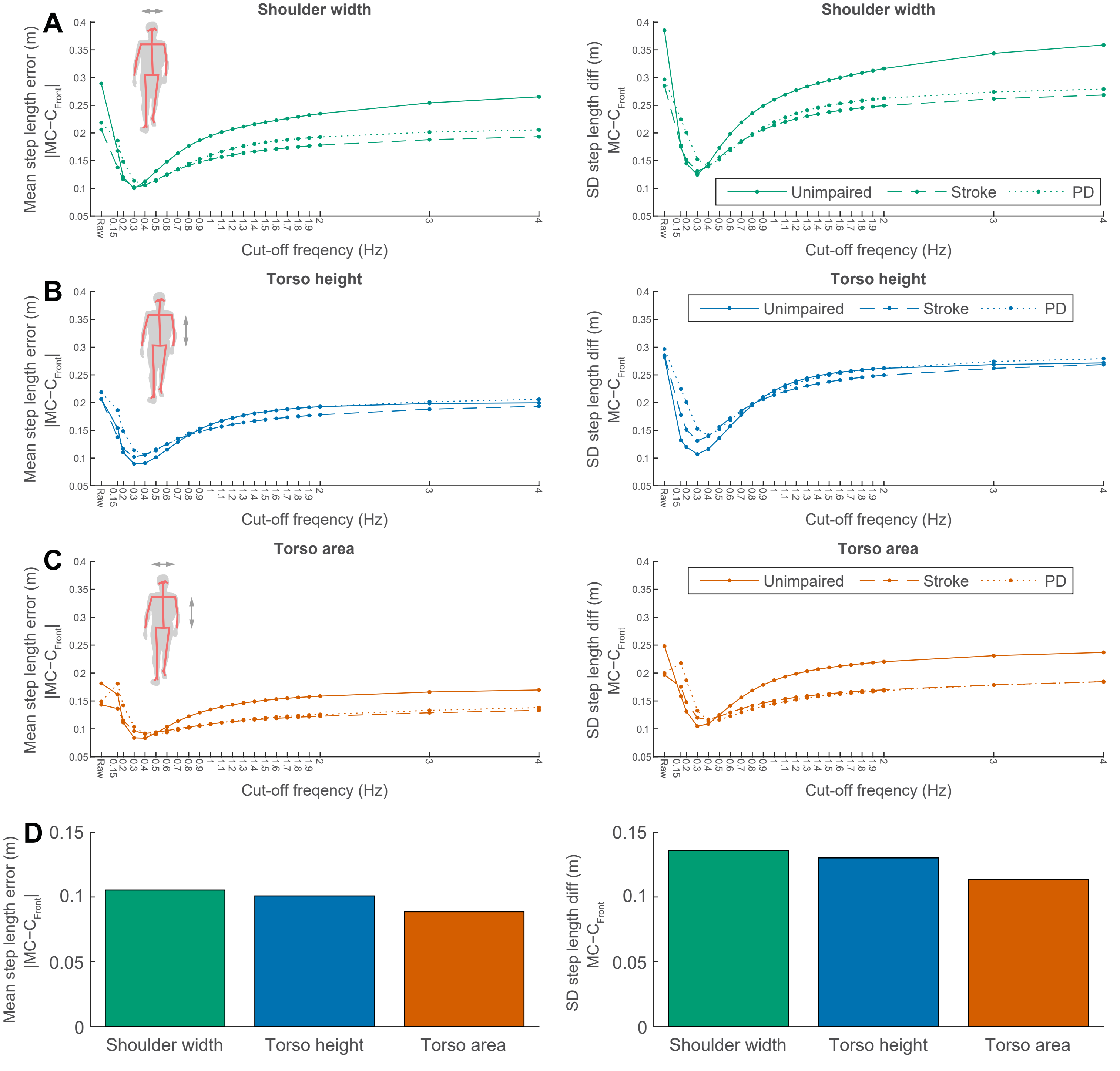


**Fig. S4. Evaluation of tracking methods and smoothing using frontal plane workflow.** We developed a frontal plane workflow that uses the pixel size of the person to calculate depth-changes throughout the walking bout (see Methods in main text). We evaluated three tracking methods: shoulder width (A), torso height (B) and torso area (C). Shoulder width is the horizontal distance between left and right shoulder keypoints, torso height is the vertical distance between the Neck and MidHip keypoints and torso area is the square root of the product of shoulder width and torso height. Furthermore, we evaluated the best smoothing method of pixel size ratios: we used raw data and low-pass filtered using cut-off frequencies ranging from 0.15 to 0.4 Hz. We calculated the mean step length error (left) and SD of step length differences (right) in order to evaluate the best method. Panel D show the performance of the tracking methods at the optimal smoothing frequency for each method (averaged across unimpaired, stroke and PD data sets). Note that photographs in panels A–C have been replaced by silhouettes to conform to medRxiv policy.

#### Fig. S5

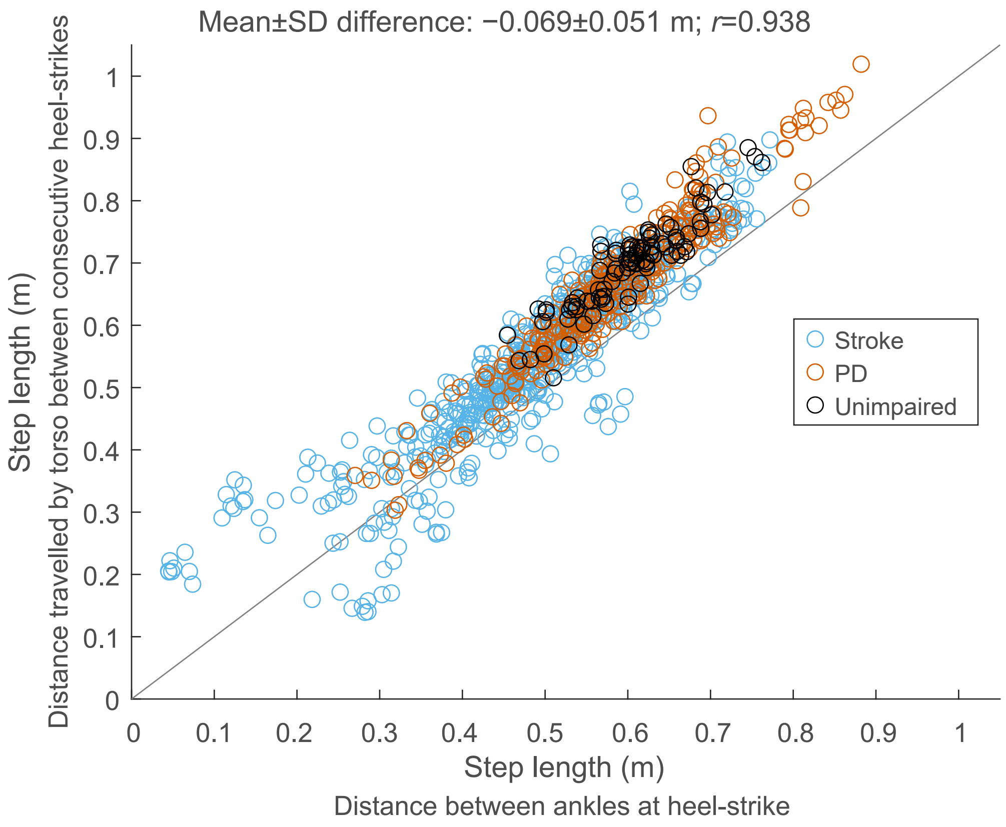


**Fig. S5. Comparison of two methods to calculate step length.** We used two methods to calculate step lengths; 1) as the horizontal distance between ankle markers or keypoints at instants of heel-strike (x-axis) and 2) as the distance travelled by the torso between consecutive bilateral heel-strikes (y-axis). We used the distance travelled by the torso because the distances between the ankles at a heel-strike instant cannot be obtained from frontal plane videos. When comparing step lengths between motion capture and sagittal plane video we used the distance between the ankles; all step length comparisons with frontal plane data used the distance travelled by the torso.

Table S1 Comparison of spatiotemporal gait parameters of the unimpaired group [a]

| Gait Parameter | Difference (Mean±SD) | | | Error (Mean±SD) | | | 95% Limits of Agreement | | |
|---|---|---|---|---|---|---|---|---|---|
| | $MC-C_{F,A}$ | $MC-C_{F,T}$ | $C_{F,A}-C_{F,T}$ | $\|MC-C_{F,A}\|$ | $\|MC-C_{F,T}\|$ | $\|C_{F,A}-C_{F,T}\|$ | $MC-C_{F,A}$ | $MC-C_{F,T}$ | $C_{F,A}-C_{F,T}$ |
| Step time (s) | | | | | | | | | |
| Step | 0.01±0.06 | −0.00±0.05 | −0.02±0.07 | 0.04±0.04 | 0.03±0.04 | 0.05±0.05 | −0.10; 0.13 | −0.10; 0.10 | −0.15; 0.11 |
| Trial | 0.01±0.02 | −0.00±0.02 | −0.02±0.02 | 0.02±0.01 | 0.01±0.01 | 0.02±0.02 | −0.02; 0.05 | −0.03; 0.03 | −0.05; 0.02 |
| Step length (m) | | | | | | | | | |
| Step | 0.020±0.124 | 0.022±0.093 | 0.001±0.156 | 0.094±0.082 | 0.074±0.060 | 0.123±0.095 | −0.222; 0.263 | −0.160; 0.203 | −0.305; 0.307 |
| Trial | 0.020±0.032 | 0.021±0.037 | 0.001±0.031 | 0.029±0.025 | 0.034±0.025 | 0.024±0.019 | −0.043; 0.084 | −0.052; 0.094 | −0.060; 0.062 |
| Gait speed ($m\ s^{-1}$) | | | | | | | | | |
| Trial | 0.01±0.06 | 0.04±0.06 | 0.04±0.06 | 0.05±0.04 | 0.06±0.05 | 0.06±0.04 | −0.11; 0.12 | −0.08; 0.17 | −0.09; 0.16 |

MC, motion capture; $C_{F,A}$, frontal plane camera that person walks away from; $C_{F,T}$, frontal plane camera that person walks toward

[a] Values of spatiotemporal gait parameters calculated as trial-level averages and for individual steps.

Table S2 Spatiotemporal gait parameters for stroke and PD groups [a]

| Gait parameter | Preferred speed (Mean±SD) | | | Fast speed (Mean±SD) | | |
|---|---|---|---|---|---|---|
| | MC | $C_S$ | $C_F$ | MC | $C_S$ | $C_F$ |
| *Stroke* | | | | | | |
| Step time (s) | | | | | | |
| Paretic | 0.77±0.31 | 0.76±0.31 | 0.74±0.32 | 0.66±0.29 | 0.65±0.29 | 0.64±0.28 |
| Non-paretic | 0.60±0.07 | 0.61±0.07 | 0.61±0.07 | 0.52±0.07 | 0.53±0.07 | 0.53±0.08 |
| Step $length_{Ankle}$ (m) [b] | | | | | | |
| Paretic | 0.480±0.094 | 0.471±0.100 | … | 0.550±0.106 | 0.538±0.113 | … |
| Non-paretic | 0.447±0.134 | 0.438±0.146 | … | 0.513±0.151 | 0.501±0.159 | … |
| Step $length_{Torso}$ (m) [c] | | | | | | |
| Paretic | 0.536±0.126 | 0.522±0.125 | 0.579±0.111 | 0.610±0.138 | 0.599±0.142 | 0.665±0.135 |
| Non-paretic | 0.510±0.130 | 0.485±0.142 | 0.527±0.168 | 0.581±0.147 | 0.564±0.160 | 0.600±0.182 |
| Gait $speed_{Ankle}$ ($m\ s^{-1}$) [b] | | | | | | |
| Trial | 0.72±0.24 | 0.71±0.25 | … | 0.98±0.35 | 0.95±0.35 | … |
| Gait $speed_{Torso}$ ($m\ s^{-1}$) [c] | | | | | | |
| Trial | 0.82±0.28 | 0.79±0.29 | 0.88±0.30 | 1.09±0.38 | 1.07±0.39 | 1.17±0.42 |
| Step time asym. | | | | | | |
| Trial | 0.09±0.13 | 0.08±0.12 | 0.07±0.13 | 0.09±0.12 | 0.07±0.11 | 0.07±0.11 |
| Step $length_{Ankle}$ asym. [b] | | | | | | |
| Trial | 0.055±0.147 | 0.058±0.159 | … | 0.055±0.153 | 0.057±0.154 | … |
| Step $length_{Torso}$ asym. [c] | | | | | | |
| Trial | 0.025±0.080 | 0.046±0.068 | 0.067±0.130 | 0.025±0.077 | 0.038±0.055 | 0.066±0.113 |
| *Parkinson's disease* | | | | | | |
| Step time (s) | | | | | | |
| Right | 0.56±0.06 | 0.56±0.06 | 0.55±0.06 | 0.47±0.06 | 0.47±0.06 | 0.46±0.06 |
| Left | 0.56±0.07 | 0.57±0.06 | 0.55±0.06 | 0.47±0.06 | 0.47±0.06 | 0.46±0.06 |
| Step $length_{Ankle}$ (m) [b] | | | | | | |
| Right | 0.527±0.094 | 0.548±0.087 | … | 0.627±0.090 | 0.643±0.081 | … |
| Left | 0.536±0.107 | 0.539±0.111 | … | 0.639±0.089 | 0.641±0.088 | … |
| Step $length_{Torso}$ (m) [c] | | | | | | |
| Right | 0.607±0.120 | 0.601±0.122 | 0.647±0.135 | 0.721±0.107 | 0.735±0.108 | 0.773±0.127 |
| Left | 0.603±0.119 | 0.620±0.123 | 0.640±0.138 | 0.715±0.104 | 0.734±0.112 | 0.792±0.162 |
| Gait $speed_{Ankle}$ ($m\ s^{-1}$) [b] | | | | | | |
| Trial | 0.95±0.19 | 0.97±0.19 | … | 1.36±0.19 | 1.38±0.19 | … |
| Gait $speed_{Torso}$ ($m\ s^{-1}$) [c] | | | | | | |
| Trial | 1.09±0.22 | 1.10±0.23 | 1.18±0.25 | 1.54±0.22 | 1.57±0.24 | 1.70±0.27 |
| Trunk incl. (°) [d] | | | | | | |
| Trial | 74.5±5.3 | 74.5±5.0 | … | 72.0±5.7 | 72.0±5.0 | … |

MC, motion capture; $C_S$, sagittal plane camera; $C_F$, frontal plane camera

[a] Values of spatiotemporal gait parameters are calculated as session-level averages.

[b] Parameter depending on step length in which step length is calculated as distance between ankles at heel-strike; missing values because step length calculated as ankle-distance cannot be calculated from $C_F$.

[c] Parameter depending on step length in which step length is calculated as distance travelled by torso between consecutive heel-strikes.

[d] Missing values because trunk inclination cannot be calculated from $C_F$.

Table S3 Comparison of spatiotemporal gait parameters of stroke and PD groups calculated as trial averages

| | Difference (Mean±SD) | | | Error (Mean±SD) | | | 95% Limits of Agreement | | |
|---|---|---|---|---|---|---|---|---|---|
| Gait Parameter | MC−$C_S$ | MC−$C_F$ | $C_F$−$C_S$ | \|MC−$C_S$\| | \|MC−$C_F$\| | \|$C_S$−$C_F$\| | MC−$C_S$ | MC−$C_F$ | $C_F$−$C_S$ |
| *Stroke* | | | | | | | | | |
| Step time (s) | | | | | | | | | |
| Away from $C_F$ [a] | 0.00±0.02 | 0.02±0.07 | 0.02±0.07 | 0.02±0.01 | 0.05±0.05 | 0.05±0.05 | −0.04; 0.04 | −0.12; 0.16 | −0.12; 0.16 |
| Toward $C_F$ | 0.00±0.03 | −0.01±0.09 | −0.01±0.09 | 0.02±0.02 | 0.06±0.07 | 0.05±0.07 | −0.06; 0.06 | −0.18; 0.17 | −0.17; 0.16 |
| Step length (m) [b] | | | | | | | | | |
| Away from $C_F$ | 0.011±0.036 | −0.056±0.090 | −0.072±0.092 | 0.028±0.026 | 0.084±0.064 | 0.091±0.073 | −0.059; 0.082 | −0.233; 0.120 | −0.252; 0.108 |
| Toward $C_F$ | 0.009±0.042 | −0.013±0.077 | −0.029±0.082 | 0.030±0.031 | 0.062±0.048 | 0.069±0.053 | −0.074; 0.092 | −0.165; 0.139 | −0.190; 0.132 |
| Gait speed (m $s^{-1}$) [b] | | | | | | | | | |
| Away from $C_F$ | 0.02±0.05 | −0.13±0.11 | −0.15±0.09 | 0.04±0.04 | 0.14±0.10 | 0.15±0.09 | −0.09; 0.12 | −0.34; 0.08 | −0.33; 0.03 |
| Toward $C_F$ | 0.02±0.09 | −0.01±0.09 | −0.04±0.10 | 0.04±0.07 | 0.06±0.06 | 0.08±0.08 | −0.15; 0.19 | −0.18; 0.16 | −0.24; 0.17 |
| Step time asym. | | | | | | | | | |
| Away from $C_F$ | 0.01±0.03 | −0.00±0.09 | −0.01±0.09 | 0.02±0.02 | 0.07±0.06 | 0.07±0.06 | −0.05; 0.07 | −0.19; 0.18 | −0.19; 0.16 |
| Toward $C_F$ | 0.02±0.04 | 0.04±0.10 | 0.03±0.09 | 0.03±0.03 | 0.08±0.08 | 0.07±0.07 | −0.06; 0.09 | −0.15; 0.24 | −0.15; 0.21 |
| Step length asym. [b] | | | | | | | | | |
| Away from $C_F$ | −0.003±0.068 | −0.064±0.143 | −0.041±0.144 | 0.047±0.049 | 0.113±0.109 | 0.107±0.103 | −0.136; 0.130 | −0.345; 0.216 | −0.322; 0.241 |
| Toward $C_F$ | −0.001±0.088 | −0.021±0.165 | −0.010±0.134 | 0.055±0.069 | 0.103±0.130 | 0.095±0.095 | −0.174; 0.172 | −0.344; 0.303 | −0.273; 0.252 |
| *Parkinson's disease* | | | | | | | | | |
| Step time (s) | | | | | | | | | |
| Away from $C_F$ | −0.00±0.01 | 0.02±0.04 | 0.02±0.04 | 0.01±0.01 | 0.03±0.03 | 0.03±0.03 | −0.03; 0.03 | −0.05; 0.09 | −0.06; 0.10 |
| Toward $C_F$ | −0.00±0.02 | 0.00±0.03 | 0.00±0.03 | 0.01±0.01 | 0.02±0.02 | 0.02±0.02 | −0.03; 0.03 | −0.06; 0.06 | −0.06; 0.06 |
| Step length (m) [b] | | | | | | | | | |
| Away from $C_F$ | −0.004±0.021 | −0.082±0.086 | −0.071±0.091 | 0.018±0.012 | 0.092±0.074 | 0.091±0.070 | −0.046; 0.038 | −0.249; 0.086 | −0.249; 0.106 |
| Toward $C_F$ | −0.017±0.024 | −0.021±0.070 | −0.009±0.072 | 0.025±0.017 | 0.055±0.048 | 0.058±0.044 | −0.065; 0.030 | −0.158; 0.117 | −0.151; 0.132 |
| Gait speed (m $s^{-1}$) [b] | | | | | | | | | |
| Away from $C_F$ | −0.01±0.04 | −0.21±0.15 | −0.19±0.15 | 0.03±0.02 | 0.23±0.13 | 0.22±0.12 | −0.08; 0.07 | −0.52; 0.09 | −0.50; 0.11 |
| Toward $C_F$ | −0.03±0.04 | −0.03±0.09 | −0.01±0.11 | 0.04±0.03 | 0.08±0.06 | 0.08±0.07 | −0.10; 0.04 | −0.22; 0.15 | −0.22; 0.20 |
| Trunk incl. (°) [c] | | | | | | | | | |
| Away from $C_F$ | −0.6±2.0 | … | … | 1.6±1.2 | … | … | −4.4; 3.3 | … | … |
| Toward $C_F$ | 0.6±1.4 | … | … | 1.2±0.8 | … | … | −2.2; 3.3 | … | … |

MC, motion capture; $C_S$, sagittal plane camera; $C_F$, frontal plane camera

[a] Values are shown for separate walking directions: 1) trials in which the person walks away from $C_F$ with their left side turned to $C_S$ or 2) trials where the person walks toward $C_F$ with their right side turned to $C_S$.

[b] Parameter depending on step length: comparisons of MC and $C_S$, step length calculated as distance between ankles at heel-strike; comparisons of MC and $C_F$ and of $C_S$ and $C_F$, step length calculated as distance travelled by torso between consecutive heel-strikes.

[c] Missing values because trunk inclination cannot be calculated from $C_F$.

Table S4 Comparison of spatiotemporal gait parameters of stroke and PD groups calculated for individual steps

| Gait Parameter | Difference (Mean±SD) | | | Error (Mean±SD) | | | 95% Limits of Agreement | | |
|---|---|---|---|---|---|---|---|---|---|
| | $MC-C_S$ | $MC-C_F$ | $C_F-C_S$ | $\|MC-C_S\|$ | $\|MC-C_F\|$ | $\|C_S-C_F\|$ | $MC-C_S$ | $MC-C_F$ | $C_F-C_S$ |
| *Stroke* | | | | | | | | | |
| Step time (s) | | | | | | | | | |
| Away from $C_F$ [a] | 0.00±0.04 | 0.02±0.12 | 0.02±0.12 | 0.03±0.02 | 0.09±0.08 | 0.09±0.08 | −0.07; 0.07 | −0.21; 0.24 | −0.21; 0.24 |
| Toward $C_F$ | 0.00±0.04 | −0.01±0.13 | −0.01±0.13 | 0.03±0.03 | 0.08±0.10 | 0.08±0.10 | −0.08; 0.08 | −0.27; 0.25 | −0.27; 0.25 |
| Step length (m) [b] | | | | | | | | | |
| Away from $C_F$ | 0.016±0.058 | −0.051±0.125 | −0.067±0.128 | 0.046±0.039 | 0.104±0.086 | 0.109±0.096 | −0.096; 0.129 | −0.296; 0.194 | −0.318; 0.184 |
| Toward $C_F$ | 0.013±0.057 | −0.012±0.103 | −0.025±0.103 | 0.045±0.036 | 0.080±0.066 | 0.083±0.066 | −0.098; 0.124 | −0.214; 0.190 | −0.227; 0.176 |
| *Parkinson's disease* | | | | | | | | | |
| Step time (s) | | | | | | | | | |
| Away from $C_F$ | −0.00±0.02 | 0.02±0.06 | 0.02±0.06 | 0.02±0.01 | 0.05±0.04 | 0.05±0.05 | −0.04; 0.04 | −0.10; 0.14 | −0.11; 0.15 |
| Toward $C_F$ | −0.00±0.02 | 0.00±0.04 | 0.00±0.05 | 0.02±0.01 | 0.03±0.03 | 0.04±0.03 | −0.05; 0.04 | −0.09; 0.09 | −0.09; 0.10 |
| Step length (m) [b] | | | | | | | | | |
| Away from $C_F$ | −0.009±0.042 | −0.070±0.125 | −0.062±0.131 | 0.035±0.026 | 0.107±0.095 | 0.109±0.095 | −0.092; 0.074 | −0.315; 0.174 | −0.318; 0.195 |
| Toward $C_F$ | −0.010±0.041 | −0.016±0.094 | −0.006±0.102 | 0.033±0.026 | 0.072±0.062 | 0.078±0.065 | −0.090; 0.070 | −0.200; 0.167 | −0.206; 0.193 |

MC, motion capture; $C_S$, sagittal plane camera; $C_F$, frontal plane camera

[a] Values are shown for separate walking directions: 1) trials in which the person walks away from $C_F$ with their left side turned to $C_S$ or 2) trials where the person walks toward $C_F$ with their right side turned to $C_S$.

[b] Parameter depending on step length: comparisons of MC and $C_S$, step length calculated as distance between ankles at heel-strike; comparisons of MC and $C_F$ and of $C_S$ and $C_F$, step length calculated as distance travelled by torso between consecutive heel-strikes.
